# Evidence of aberrant anti-Epstein-Barr virus antibody response, though no viral reactivation, in people with post-stroke fatigue

**DOI:** 10.1101/2024.04.08.24305483

**Authors:** Isobel C. Mouat, Judy Zhu, Alperen Aslan, Barry W. McColl, Stuart M. Allan, Craig J. Smith, Marion S. Buckwalter, Laura McCulloch

## Abstract

**BACKGROUND:** Fatigue is a common complication of stroke that has a significant impact on quality of life. The biological mechanisms that underly post-stroke fatigue are currently unclear, however, reactivation of latent viruses and their impact on systemic immune function have been increasingly reported in other conditions where fatigue is a predominant symptom. Epstein-Barr virus (EBV) in particular has been associated with fatigue, including in long-COVID and myalgic encephalomyelitis/chronic fatigue syndrome, but has not yet been explored within the context of stroke.

**AIMS:** We performed an exploratory analysis to determine if there is evidence of a relationship between EBV reactivation and post-stroke fatigue.

**METHODS:** In a chronic ischemic stroke cohort (>6 months post-stroke), we assayed circulating EBV by qPCR and measured the titres of anti-EBV antibodies by ELISA in patients with high fatigue (FACIT-F <40) and low fatigue (FACIT-F >41). Statistical analysis between two-groups were performed by t-test when normally distributed according to the Shapiro-Wilk test, by Mann-Whitney test when the data was not normally distributed, and by Fisher’s exact test for categorical data.

**RESULTS:** We observed a similar incidence of viral reactivation between people with low versus high levels of post-stroke fatigue (5 of 22 participants (24%) versus 6 of 22 participants (27%)). Although the amount of circulating EBV was similar between the groups, we observed an altered antibody response against EBV antigens in participants with high fatigue, with reduced IgM against the Viral Capsid Antigen (VCA; 2.244 ± 0.926 vs 3.334 ± 2.68; P = 0.031). Total IgM levels were not different between groups indicating this effect was specific to EBV (3.23 × 10^5^ ± 4.44 × 10^4^ high fatigue versus 4.60 × 10^5^ ± 9.28 × 10^4^ low fatigue; P= 0.288).

**CONCLUSIONS:** These findings indicate that EBV is not more prone to reactivation during chronic stroke recovery in those with post-stroke fatigue. However, the dysregulated antibody response to EBV may be suggestive of viral reactivation at an earlier stage after stroke and warrants further investigation.

## Introduction

Post-stroke fatigue is a major clinical concern that is experienced by around half of stroke survivors (1). Fatigue has a major impact on quality of life and has been identified by stroke survivors, carers and health care professionals as a high priority for future research (2). There are varying estimates of the duration of post-stroke fatigue, though it is clear that for many it is a persistent symptom that continues for years (3). Multiple factors are hypothesised to contribute to risk of post-stroke fatigue, including biological sex, white matter abnormalities, stroke severity, and depression (4). Despite its clinical importance, there are currently no effective treatments, and identifying biological mechanisms driving post-stroke fatigue has been recommended as a research priority, with dysregulation of the immune system and inflammation highlighted as the top area for further investigation (5).

With the recent worldwide impact of long-COVID, there has been an upsurge of interest in understanding common factors that drive fatigue in different clinical conditions. Reactivation of Epstein-Barr virus (EBV) and its association with a dysregulated immune system has been increasingly associated with fatigue, including in long-COVID (6) and myalgic encephalomyelitis/chronic fatigue syndrome (ME/CFS) (7). EBV is a ubiquitous gamma-herpesvirus virus that infects >90% of all people (8). EBV lies dormant for the majority of life, and constant host immune surveillance largely maintains the virus in a latent state. There can be however periodic reactivation, particularly during periods of immunosuppression (9). EBV reactivation has been observed in an array of conditions where the immune system is compromised, including haematopoietic and solid organ transplantation (10, 11), HIV infection (12, 13), and stress (14). While it is still unknown how EBV is linked to the development of fatigue, it is thought that genetic susceptibility (15) and viral-induced chronic inflammation (16) may both have a role.

Suppression of some aspects of systemic immune function is a common feature of stroke and is associated with infection vulnerability and worse outcomes (17-20). While alterations to peripheral immunity are most apparent in the acute phase of stroke recovery, there is evidence of persistent immune changes during chronic recovery (21). Therefore, post-stroke immune suppression has potential to create an immune environment permissible to reactivation of EBV. There is some evidence of herpesvirus reactivation following stroke: in the five years following stroke, there is an increased risk of herpes zoster reactivation in people (22). Animal models indicate risk of both HIV and herpes simplex virus 1 (HSV-1) reactivation following stroke (23, 24). We focused on EBV due to its high seroprevalence. EBV also has the ability to modulate immune function, including suppression of IFNγ and CD8 T cell responses, and the prevention of B cell apoptosis, resulting in increased risk of cancer and autoimmune disease (25-28), suggesting reactivation of EBV after stroke could contribute to further dysregulation of the immune environment and development of secondary complications.

In this study we aimed to investigate the frequency of circulating EBV and the immunoglobulin response to EBV in people recovering from stroke with high versus low levels of reported fatigue.

## Methods

### Study design and participants

The StrokeCog study is a longitudinal cohort study aiming to investigate the post-stroke immune response and cognitive functioning over the course of stroke recovery (29). Participants included in the parent study had ischemic strokes that were confirmed by MRI or CT scan, were over the age of 25, were able to return for annual follow-up visits, and had fluency in English. Exclusion criteria included preceding cognitive impairment or dysphasia that precluded ability to complete the neurological assessments, life expectancy of under one year, or pre-existing conditions that would impact the assessment of neurological or cognitive outcomes. The parent study was approved by the Stanford University Institutional Review Board. We designed this as a nested case-control (fatigue vs. no fatigue) cohort within the StrokeCog cohort. Cohort characteristics are further discussed in the results section.

### Clinical data

A neurological evaluation was performed on each participant comprised of various neuropsychological assessments (29). For the purpose of this study, we are particularly interested in fatigue, mood, and cognition. Fatigue was measured in participants using the Functional Assessment of Chronic Illness Therapy – Fatigue Scale (FACIT-F) (30), a questionnaire tool that assesses patient-reported fatigue and impact on daily living. The measure is made up of 13 questions, each requiring an answer on a 5-point Likert-type scale, with the summation of responses producing a single value. Additionally, the Stroke Impact Scale (SIS) version 3 was administered, a well-established patient-based, self-report questionnaire (31). Various domains are measured, and for the purpose of this study we examined the SIS3 domain, which is a measure of mood/emotion. Finally, to measure cognition, the Montreal Cognitive Assessment (MoCA) was utilized (32).

### Sample collection and processing

Blood was collected the same time as neurological assessment. For the isolation of plasma and peripheral blood mononuclear cells (PBMCs), blood was collected into sterile BD Vacutainer tubes containing 15% K3 ethylenediaminetetraacetic acid (EDTA) solution (Thermo Fisher Scientific), placed on ice, and centrifuged at 2,000g for 10 minutes at 4°C. Plasma supernatant was isolated, aliquoted, and stored at -80°C prior to use. The remaining sample was further processed with a 15mL Ficoll density gradient in a SepMate Tube-50, cells were washed with PBS with 2% fetal bovine serum (FBS), counted on a haemocytometer, and frozen in a Mr. Frosty (Thermo Fisher) at -80°C for 24 hours before being transferred to liquid nitrogen for storage. Genomic DNA was extracted from 2x10^6^ PBMCs with the Qiagen QIAamp DNA Blood Mini Kit (cat no 51104).

### Detection of antibodies

Enzyme-linked immunosorbent assay (ELISA) was used to measure antibody titres in plasma including those against EBV antigens and C-reactive protein (CRP) and total IgM and IgG. Commercial ELISA kits were used to measure anti-viral capsid antigen (VCA) IgM (Abcam, ab108732), anti-VCA IgG (Abcam, ab108730), anti-Epstein-Barr nuclear antigen 1 (EBNA1) IgG (Abcam, ab108731), IgM (Abcam, ab214568), IgG (Abcam, ab195215), and anti-CRP (Abcam, ab260058), and completed according to manufacturer’s instructions. Plasma samples were diluted to 1:100 concentration for the anti-VCA IgM and anti-EBNA1 IgG, 1:2000 for the anti-VCA IgG and anti-CRP and 1:100,000 for the IgM and total IgG. Samples were run in duplicate and each plate was run with positive, negative, and cut-off controls (anti-VCA and anti-EBNA1), or a standard. Absorbance was measured at 450 nm using a SpectraMax 340PC384 (Molecular Devices). Interpretation of results was done in accordance with manufacturer’s instructions. For anti-EBV titres, samples were considered to give a positive result if more than 10% above the cut-off control.

### EBV viraemia

Circulating EBV was measured by qPCR with gDNA isolated from peripheral leukocytes. The Norgen EBV TaqMan PCR kit (TM41050) was used and duplicate samples were run on a BioRad CFX96 Real-Time PCR Detection System (BioRad). The assay was completed and results were analysed in accordance with manufacturer’s instructions. All samples displayed amplification of the internal PCR validation control.

### Statistical analyses

Analysis and presentation of data was performed with GraphPad Prism software version 9.5.1 (GraphPad Software Inc.). Statistical analysis between two-groups were performed by t-test when normally distributed according to the Shapiro-Wilk test, by Mann-Whitney test when the data was not normally distributed, and by Fisher’s exact test for categorical data. Correlation analyses were performed by simple linear regression on log-transformed data, with best-fit line and 95% confidence bands plotted. P < 0.05 was considered statistically significant.

## Results

### Cohort selection

A total of 44 participants from the StrokeCog cohort were included in this exploratory study based on their experience of fatigue. Patients completed a fatigue questionnaire alongside a battery of neurological tests at a minimum of five months post stroke onset and a concurrent blood sample was taken. To identify participants for this study we selected the 30 individuals with the highest FACIT-F scores and 30 participants with the lowest FACIT-F scores, and of those identified 22 participants in each group that matched with regards to age, biological sex, infarct volume, initial stroke severity (National Institutes of Health Stroke Scale, NIHSS), and time from stroke to sample collection (Table 2). The resulting cohort comprised of a high fatigue (FACIT-F <40) and low fatigue (FACIT-F >41) group (Figure 1A), with the dichotomisation aligning with previously published values for levels of post-stroke fatigue (33). The higher fatigue group had lower Stroke Impact Scale domain 3 (SIS3) scores, indicating worse mood, and there was association between fatigue (FACIT-F) and mood (SIS3) (Figure 1B, C). The cohort was comprised of individuals with relatively low severity strokes, with a median NIHSS of 4 and 5 for the low and high fatigue groups, respectively, and a median infarct volume of 7.2mL (low fatigue) and 5.5mL (high fatigue, Table 1). The low and high fatigue groups had similar prevalence of risk factors, including hypertension and previous stroke, as well as medication usage. However, there was a small, but not significant, increase in duration from index stroke in the high fatigue group (Table 1).

**Table 2.** Clinical cohort. Median and interquartile range (IQR), analysed by Mann-Whitney test for nonparametric data and Fisher’s exact test for categorical data. NIHSS, National Institutes of Health Stroke Scale; IV, intravenous; IA, intra-arterial; ACE, Angiotensin Converting Enzyme.

|  | High fatigue | Low fatigue |
| --- | --- | --- |
| **Age at Baseline, median (IQR)** | 65 (56-71) | 66 (58-71) |
| **Sex** |  |  |
| Male, n (%) | 14 (64) | 14 (64) |
| Female, n (%) | 8 (36) | 8 (36) |
| **Presentation** |  |  |
| Right hemisphere, n (%) | 13 (59%) | 10 (45%) |
| Left hemisphere, n (%) | 9 (41%) | 10 (45%) |
| Bilateral, n (%) | 0 (0%) | 2 (9%) |
| NIHSS, median (IQR) | 5 (2-11) | 4 (2-13) |
| Infarct volume (mL), median (IQR) | 5.5 (0.46-39.08) | 7.2 (1.98-42.33) |
| Days from Stroke to Baseline, median (IQR) | 316 (235-1062) | 256 (206-1236) |
| IV thrombolysis, n (%) | 4 (20%) | 8 (38%) |
| IA thrombectomy, n (%) | 4 (20%) | 6 (29%) |
| **Risk Factors** |  |  |
| Hypertension, n (%) | 15 (75%) | 9 (43%) |
| Previous stroke, n (%) | 4 (24%) | 2 (12%) |
| Atrial fibrillation, n (%) | 3 (17%) | 5 (24%) |
| Hyperlipidaemia, n (%) | 9 (53%) | 6 (29%) |
| Other cardiovascular diseases, n (%) | 2 (12%) | 3 (17%) |
| Heart valve repair/replacement, n (%) | 1 (5%) | 2 (9%) |
| Angina, n (%) | 0 (0%) | 0 (%) |
| Congestive heart failure, n (%) | 1 (5%) | 1 (5%) |
| Pacemaker and/or defibrillator, n (%) | 0 (0%) | 2 (9%) |
| Coronary angioplasty, n (%) | 3 (15%) | 4 (18%) |
| **Medications** |  |  |
| Antiplatelets, n (%) | 6 (27%) | 5 (23%) |
| Anticoagulants, n (%) | 1 (5%) | 2 (9%) |
| Statin, n (%) | 18 (82%) | 16 (73%) |
| Beta-blocker, n (%) | 6 (27%) | 6 (27%) |
| ACE-inhibitor, n (%) | 4 (18%) | 4 (18%) |
| Angiotensin II receptor blocker, n (%) | 6 (27%) | 4 (18%) |

**Figure 1.**
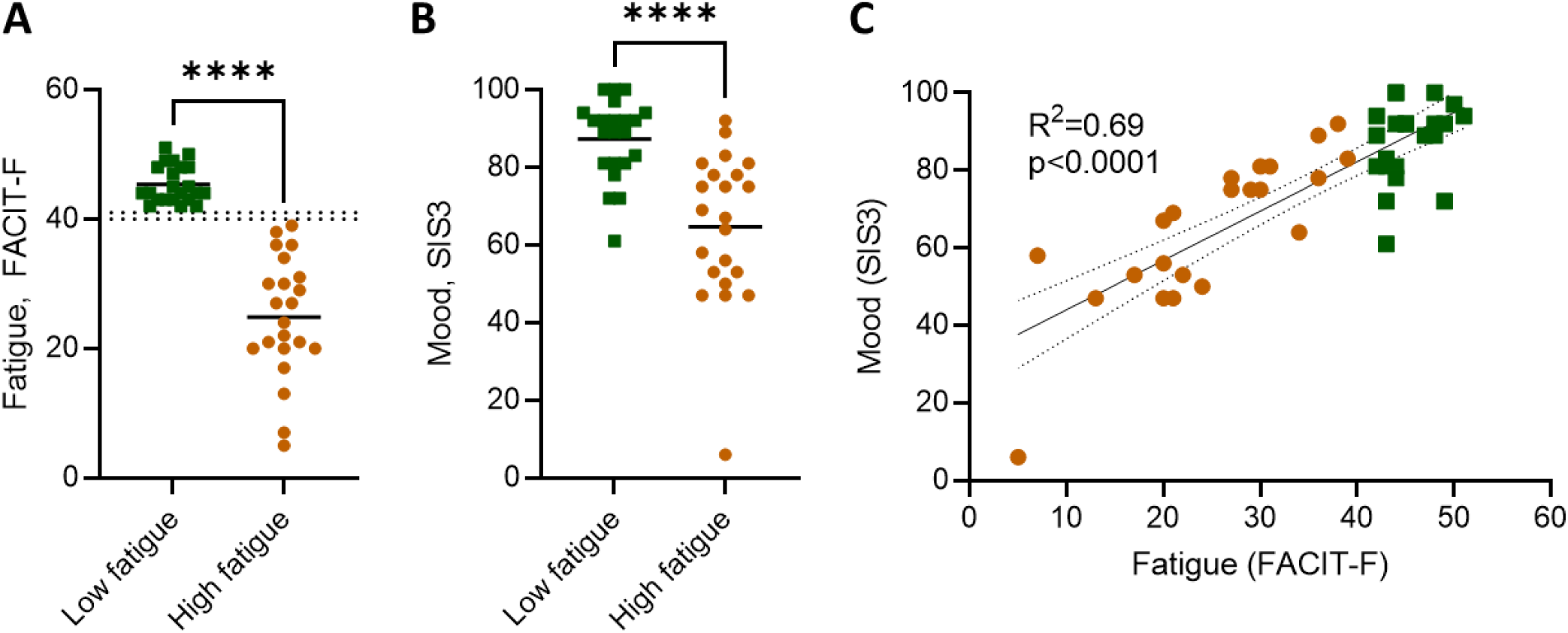
Fatigue and mood of cohort. (A) Fatigue, measured by the Functional Assessment of Chronic Illness Therapy – Fatigue (FACIT-F) tool, with threshold for high versus low fatigue demarcated by dotted lines (B) mood, as measured by the Stroke Impact Scale (SIS), and correlation between fatigue (FACIT-F) and mood (SIS3). Each data point represents an individual participant, mean plotted. Data show individual participants plus mean; analysed by (A, B) Mann-Whitney test and (C) linear regression, with best-fit line and 95% confidence band plotted; ****P < 0.0001.

### Frequency of circulating virus is not different between participants with high and low post-stroke fatigue

EBV is expected to be detectable in circulation during primary infection and reactivation events, though not during latency when the virus lays dormant in lymphoid tissues. EBV DNA was detected in the circulation of 5 of 22 participants (24%) with low fatigue and 6 of 22 participants (27%) with high fatigue (Figure A). Of the individuals positive for EBV, no difference was observed in the viral load between those with low and high fatigue (Figure B). As immune suppression after stroke is associated with increased stroke severity, we would expect that EBV reactivation would be more prevalent in participants with a higher stroke severity. Between participants with and without detectable circulating EBV, there was no difference in the initial stroke severity, measured by NIHSS (Figure 2C).

**Figure 2.**
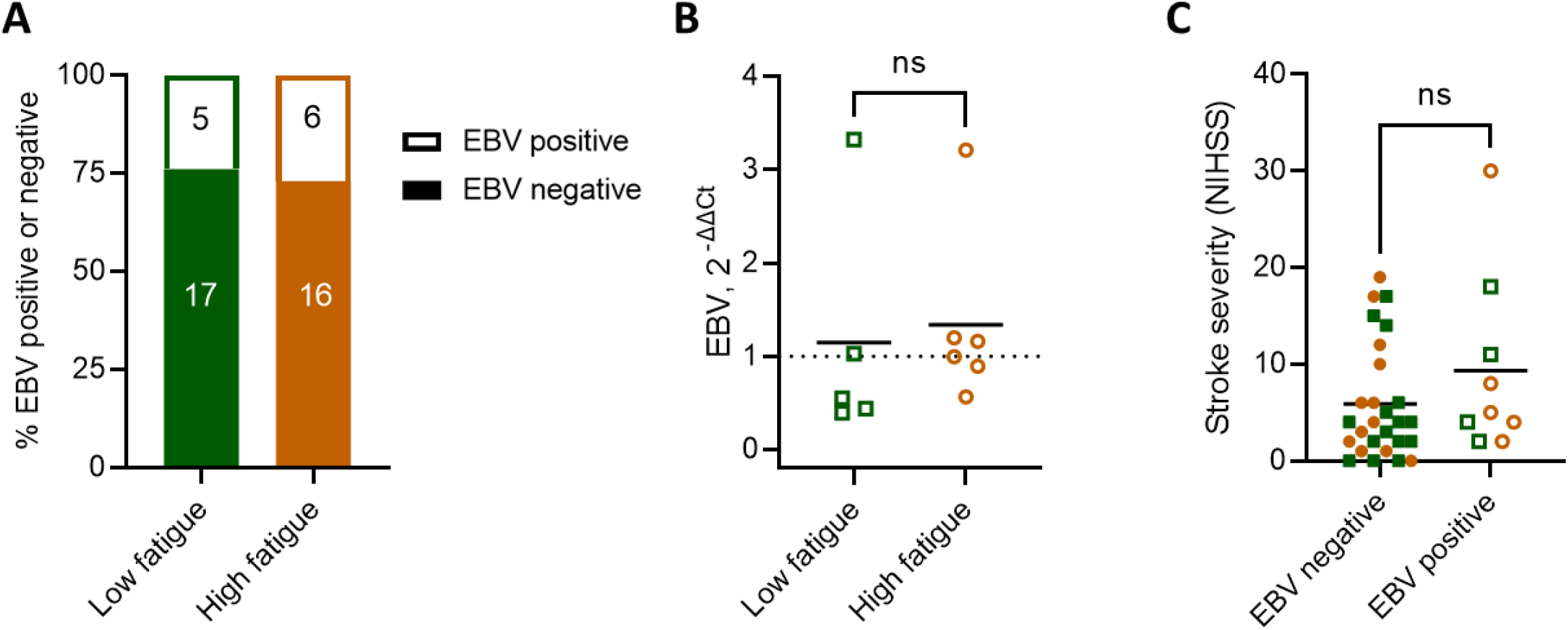
Similar frequencies of EBV viraemia in people with and without post-stroke fatigue. (A) Circulating EBV was measured by qPCR. (B) Circulating EBV load was assessed in the subset of participants positive for EBV. EBV was quantified with the delta-delta Ct method, using the housekeeping gene provided by the Norgen EBV PCR kit. (C) Stroke severity, measured by NIHSS, between participants with and without detectable circulating EBV by qPCR. Low fatigue participants are plotted as green squares and high fatigue as orange circles. Open shapes indicate participants with EBV viremia, as measured by qPCR. Data show individual participants plus mean; (B) unpaired t-test; (C) Mann-Whitney test.

### EBV seroresponse in people with post-stroke fatigue

To investigate the antibody response to EBV, we measured circulating antibodies against the EBV lytic antigen VCA and latency antigen EBNA1. We observed that all participants were seropositive for EBV, as measured by the presence of at least one IgG antibody class against EBNA1, IgG against VCA, or IgM against VCA. No recent primary infections were detected, defined as a positive IgM anti-VCA and negative IgG anti-EBNA1 (data not shown). This means that all patients showing positive viremia by qPCR were experiencing reactivation of previously latent infection.

We first measured IgM against VCA, which would be expected to be highest during primary infection and remain persistently detectable in seropositive individuals (Figure 3A). While nearly all samples were below the cut-off control, indicating a lack of recent primary infection, all samples were in the detectable range of the assay. We observed a lower IgM titre against VCA in the high fatigue group, compared to the low fatigue group (Figure 3B). Next, we measured the titre of IgG against VCA, which would be expected to increase in response to viral reactivation (Figure 3A). We did not observe a difference in anti-VCA IgG titre between groups (Figure 3C). This result aligns with the qPCR results where patients with viremia were evenly distributed between low and high fatigue groups, and reinforce that people with higher levels of post-stroke fatigue did not show increased incidence of EBV reactivation following stroke. Individuals with viremia detected by qPCR had higher titres of anti-VCA IgG (Figure 3D), as expected in response to an ongoing infection (Figure 3A).

**Figure 3.**
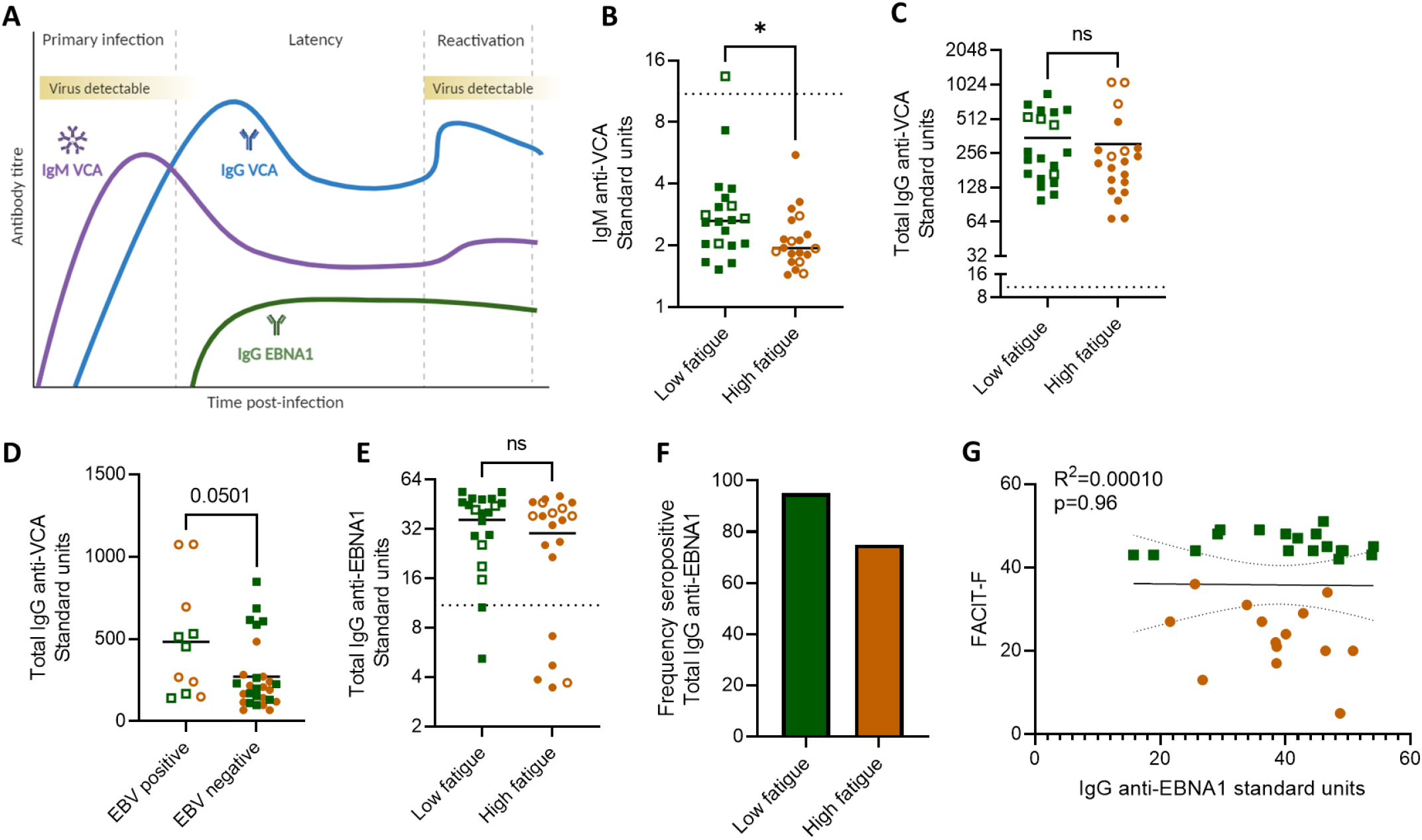
Anti-EBV seroresponse in people with post-stroke fatigue. (A) Schematic of expected circulating anti-EBV antibody titres following primary infection, and during latency, and reactivation. Created with BioRender. (B-G) Plasma antibodies to EBV were measured by ELISA. Low fatigue participants are plotted as green squares and high fatigue as orange circles. Open shapes indicate participants with EBV viremia, as measured by qPCR, and the dotted line (B, E, G) indicates the ELISA cut off control value. (B) Titre of anti-Epstein-Barr nuclear antigen 1 (EBNA1) IgG between participants with low versus high post-stroke fatigue. (C) Percent of participants within each group that are positive seropositive for anti-EBNA1 IgG. Seropositivity is considered any value >10% over the cut-off control value. (D) Association between self-reported fatigue (FACIT-F) and IgG anti-EBNA1 titre. (E) Titre of IgG anti-viral capsid antigen (VCA) between low and high fatigue groups. (F) Titre of IgG anti-VCA between participants with and without EBV viremia, measured by qPCR. (G) Titre of IgM anti-viral capsid antigen (VCA) between low and high fatigue groups. Data show individual data points plus mean; *p < 0.05; (E) unpaired t-test (B, F, G) Mann-Whitney test; (D) linear regression, with best-fit line and 95% confidence band plotted.

We next measured the titres of IgG against latency antigen EBNA1, indicative of memory to primary infection and/or an adaptive antibody response to reactivation, and did not observe a difference between low and high post-stroke fatigue groups (Figure 3E). The ELISA cut off control was used to determine seropositivity, i.e. the presence of antibody responses to antigen. Seropositivity was then used to stratify participants into seropositive versus seronegative for IgG against EBNA1. We observed that of the low fatigue participants, 18/20 (90%) displayed seropositivity for anti-EBNA1 IgG, compared to 15/20 (75%) of the high fatigue participants (Figure 3F). Of the individuals seropositive for anti-EBNA1 IgG, we did not observe a relationship between IgG anti-EBNA1 titres and fatigue (Figure 3G). Taken together with our previous finding that individuals with high fatigue have lower IgM titres against the lytic VCA antigen (Figure 3B), this could indicate a potentially aberrant antibody response to EBV in people with post-stroke fatigue. Titres of anti-EBV antibodies did not associate with initial stroke severity (Supplementary Figure 1).

We then examined anti-VCA IgM titres in relation to various clinical characteristics. As expected from our previous results (Figure 3), we observed a correlation between fatigue and anti-VCA IgM titre (Figure 4A). No association was observed between IgM against VCA and age, biological sex, days from stroke to baseline, mood, or infarct volume (Figure 4B-F). Finally, anti-VCA IgM was not associated with cognitive domains, including working memory, processing speed, memory, visual spatial, or language (Supplementary figure 2).

**Figure 4.**
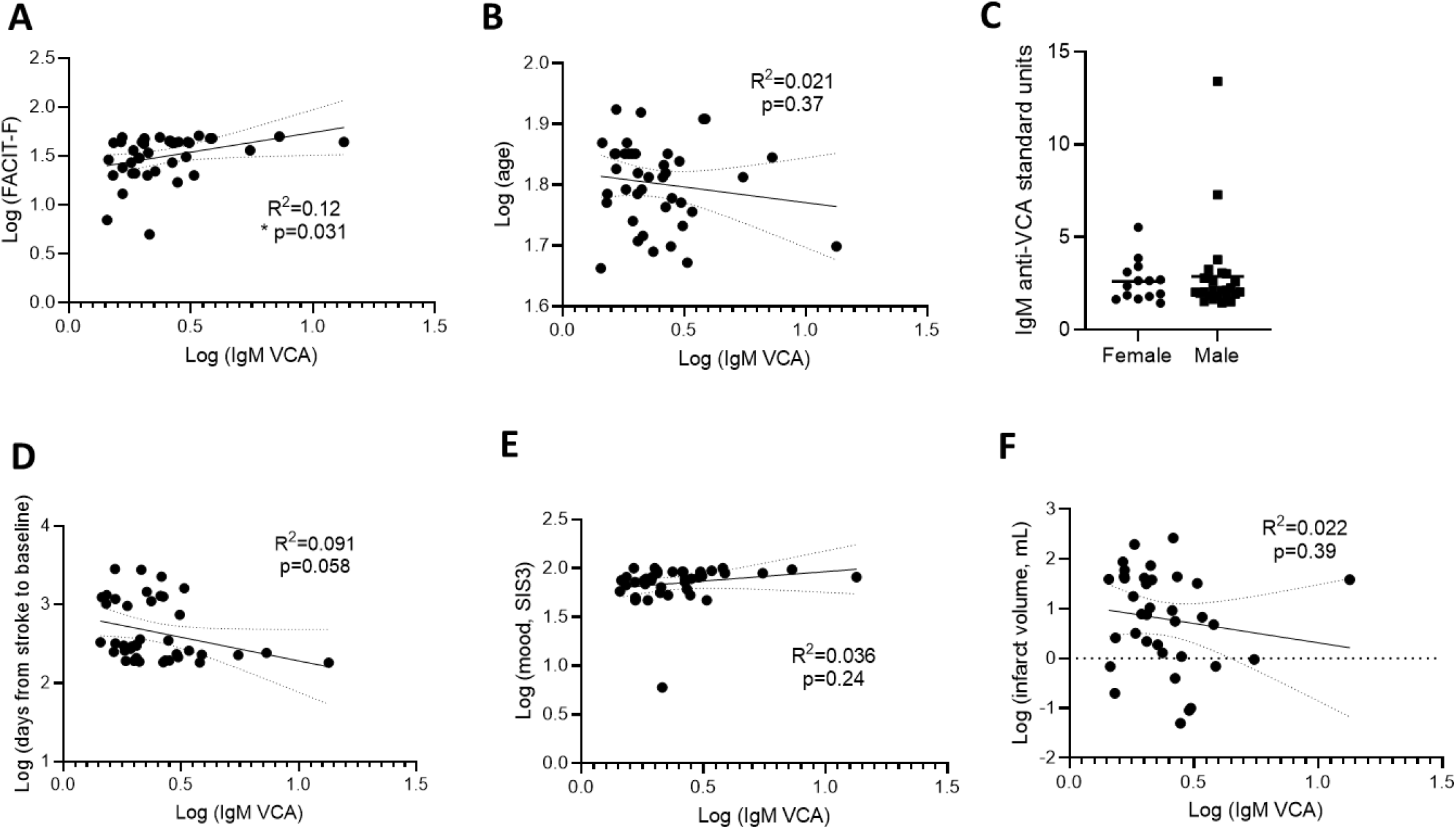
IgM anti-VCA titre correlates with fatigue but not selected clinical characteristics. Titre of IgM against VCA were plotted against various characteristics, including (A) FACIT-F score, (B) age, (C) biological sex, (D) days from stroke to baseline, (E) mood as measured by SIS3, and (G) infarct volume. Each data point represents an individual participant. Analysed by linear regression with best-fit line and 95% confidence band plotted (A-B, D-F).

Together, these results indicate that there is not a difference in EBV reactivation between groups, but suggest possible alterations to the EBV antibody response, particularly the anti-VCA IgM response, in people with post-stroke fatigue. The dampened IgM against VCA observed in participants with post-stroke fatigue does not associate with other clinical or neurological parameters, suggesting a link to fatigue in particular.

### Total antibody titres unchanged between low and high post-stroke fatigue groups

To investigate if changes to total circulating antibody levels were responsible for the observed changes in anti-EBV antibodies, we measured circulating IgM and total IgG. We did not observe differences in the levels of IgM nor total IgG between the low and high fatigue groups (Figure 5A, B), indicating that the observed differences in anti-VCA IgM were not driven by a loss of total circulating IgM.

**Figure 5.**
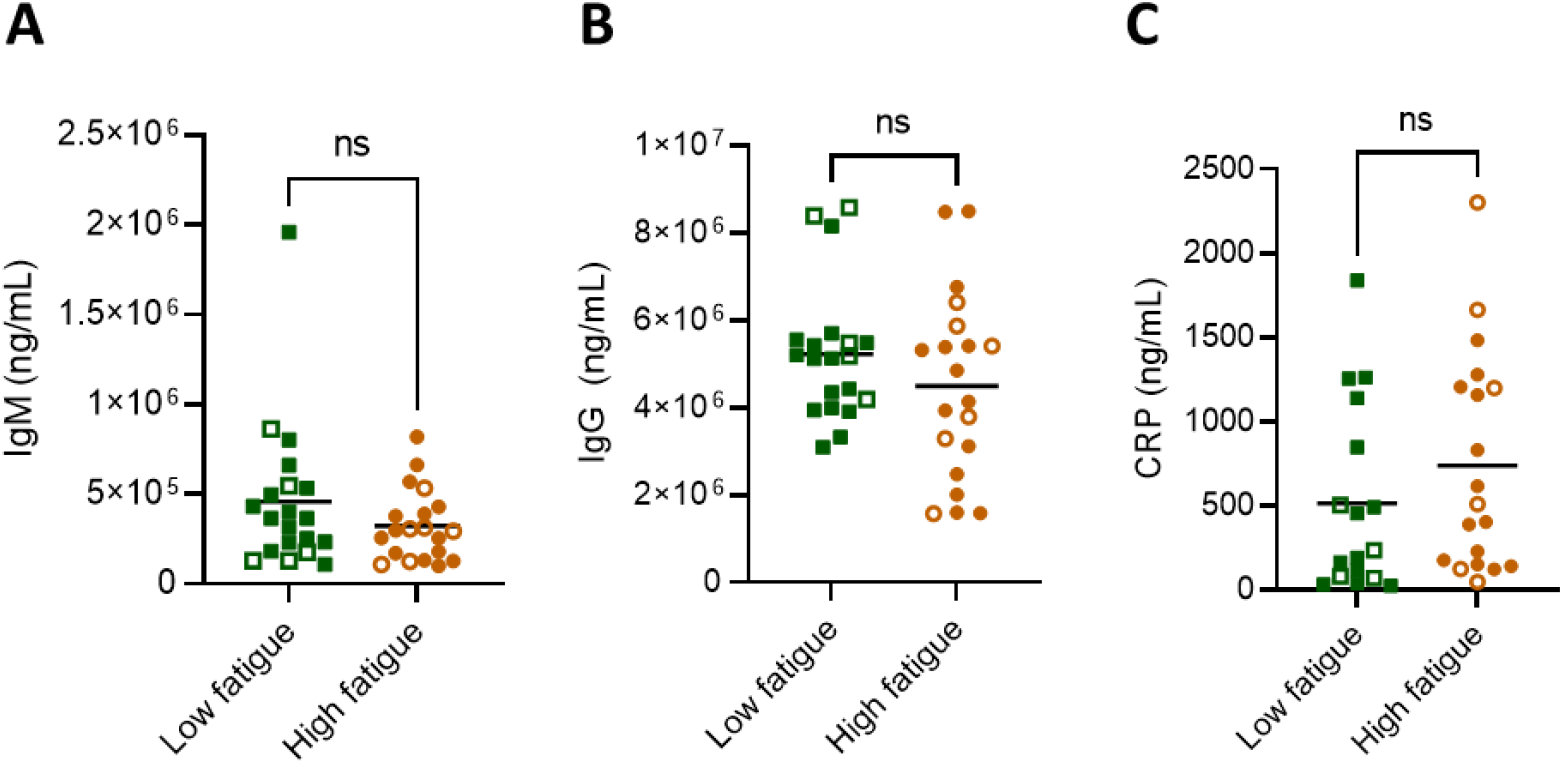
Total circulating antibodies unchanged between low and high post-stroke fatigue groups. Amount of circulating IgM and total IgG antibodies, as well as C-reactive protein (CRP). Data show individual participants plus mean; analysed by Mann-Whitney test.

Additionally, we measured the inflammatory protein C-reactive protein (CRP) that has been previously associated with poor outcome after stroke – and is indicative of stroke-induced systemic immune alterations (34). We did not observe a difference in the level of circulating CRP between those with low and high post-stroke fatigue, suggesting that fatigue is not associated with general systemic immune alterations post-stroke (Figure 5C).

No differences in antibody or CRP levels were observed in the participants with EBV viremia (open shapes) compared to those without detectable circulating virus (filled shapes, Figure 5A-C).

## Discussion

In this study, we aimed to explore if EBV reactivation associates with post-stroke fatigue. To our knowledge, this is the first study to examine EBV, and its relationship with fatigue, during stroke recovery. Here, we report a lack of evidence for EBV reactivation in people with post-stroke fatigue during chronic stroke recovery. However, we did see alterations in the antibody response to EBV in patients with high fatigue. There were no differences in total IgM or IgG antibody concentrations between high and low fatigue groups, suggesting this is specifically associated with the immune response to EBV.

Estimates of EBV reactivation in the general population vary but have been reported between 30 and 50% (35-37), and reactivation is known to increase with age (38). In this study, we detected EBV in 25% of participants, indicating that people with a history of stroke may not have elevated EBV reactivation, although a direct comparison with healthy participants would be necessary to confirm. Severe COVID-19 is known to drive similar immunological changes seen after stroke, including lymphopenia (39). In chronic recovery from COVID-19, increased IgG titres against EBV lytic antigen were shown to be a common feature in patients who experienced long-COVID, associated with high fatigue, versus those who experienced a healthy recovery (6). A smaller study confirmed reactivation of EBV in 50% of patients with long-COVID versus 20% with healthy recovery (40). However, a study of patients admitted to the intensive care unit with ongoing severe COVID-19 measured EBV reactivation in 82% of patients, which may suggest peak viral reactivation occurs doing acute illness (41). This is supported by a study looking at the sequalae of long-COVID development showing that viral reactivation occurs during acute infection and is a predictor of long-COVID (42). Together, these studies suggest that time post infection is an important consideration when investigating EBV reactivation, but the immunological effects of this may persist beyond EBV reactivation itself.

We examined EBV reactivation by measuring circulating virus and anti-EBV antibody titres and by both measures there was no evidence of EBV reactivation. Surprisingly, even in the absence of reactivation, there were indications of an aberrant anti-EBV immune response in participants with post-stroke fatigue. The IgM titre against lytic EBV antigen, VCA, was decreased in participants with higher post-stroke fatigue and was also negatively correlated with post-stroke fatigue. Additionally, the IgG titre against latency antigen EBNA1 was seropositive in a lower proportion of high fatigue participants, however we did not observe a correlation with fatigue. The mechanism and clinical relevance of this unexpected finding is not clear from current data. Stroke induces a systemic suppression of immune response, the extent of which is associated with the initial stroke severity (43). We have previously shown that B cell numbers, and antibody responses, are reduced after stroke (19, 44). Therefore, it could be hypothesised that the altered antibody response to EBV in patients with high fatigue is due to higher initial stroke severities and more extensive alterations to systemic immune function. However, we saw no association between titres of anti-VCA IgM and infarct volume or stroke severity. Furthermore, there were no differences in total IgM or in CRP concentrations between low and high fatigue groups. This suggests that the altered antibody titres to EBV antigens is not simply a result of more extensive systemic immune alterations in the high fatigue patient group. We chose to examine patients at least five months (and up to eight years) post-stroke as fatigue is a chronic condition and we aimed to examine if active EBV infection may drive fatigue. However, we know that immune alterations that occur acutely after stroke are associated with increased infection risk. Therefore, it is possible that EBV reactivation could occur more acutely after stroke. The reduced antibody titres to EBV antigen in chronic recovery in high fatigue patients may be an indication of an aberrant immune response to earlier viral reactivation.

In addition to sampling time point, there are further limitations to consider in this study. The sample size is relatively small. Additionally, fatigue is a subjective measure, reliant on patient reporting. In this study fatigue was examined by the FACIT-F tool, however various tools exist to measure fatigue and there can be discrepancies in the extent of fatigue recorded depending on which test is used (45). Here we examined only EBV, though there is evidence that other viruses reactivate following stroke, such as herpes zoster virus (22). Reactivation of multiple viruses, including cytomegalovirus and human herpesvirus (HHV) 6, HHV-7, HHV-8 have been associated with chronic fatigue and could play a role in fatigue after stroke (46).

Although we have not found an association with EBV reactivation during chronic stroke recovery and incidence of fatigue, the alterations to circulating EBV antibody titres suggest we should not fully dismiss a potential role for reactivation of EBV and/or other latent viruses in the pathophysiology of post-stroke fatigue. Further, larger studies using acute recovery timepoints and alternative latent viruses will be required to fully examine the role of viral reactivation in post-stroke fatigue.

## Data Availability

All data produced in the present study are available upon reasonable request to the authors

## FIGURES AND TABLES

Supplementary Figure 1. Anti-EBV antibodies do not associate with stroke severity.

Supplementary Figure 2. IgM anti-VCA titre does not associate with various cognitive domains

## Funding acknowledgements

This research was funded by the Leducq Stroke-IMPaCT Transatlantic Network of Excellence (19CVD01; MSB/ICM/BM/SA/CS), an American Heart Institute / Allen Frontiers Group Brain Health Award (19PABHI34580007; MSB), the Wu Tsai Neurosciences Institute (MSB), and a Wellcome Sir Henry Dale Fellowship (220755/Z/20/Z; LM).

## Author contributions

ICM, BM, SA, CS, MSB, and LM contributed to the concept and design of the work. ICM, JZ, and AA performed data acquisition. ICM completed data analysis and drafted the article. ICM, MSB, and LM contributed to interpretation of data. BM, SA, CS, MSB, and LM provided critical revision.

## Open Access

For the purpose of open access, the author has applied a CC BY public copyright licence to any Author Accepted Manuscript version arising from this submission.

## Data availability

The raw and anonymized data used in this study can be made available to other researchers on request. Written proposals can be addressed to the corresponding author and will be assessed by the StrokeCog investigators for appropriateness of use, and a data sharing agreement will be put in place before data are shared.

