## Supplemental data for "Evidence of aberrant anti-Epstein-Barr virus antibody response, though no viral reactivation, in people with post-stroke fatigue"

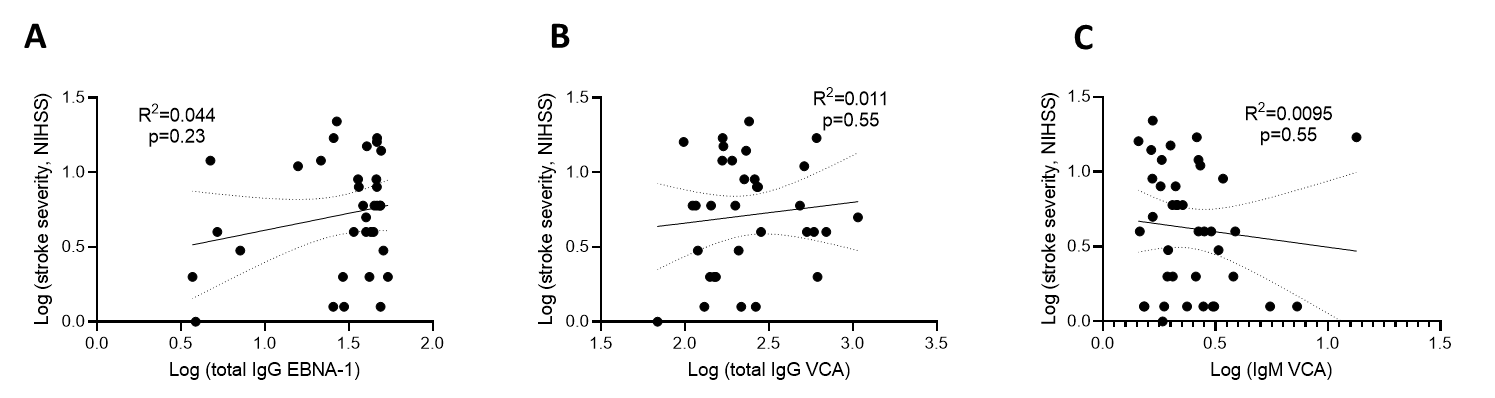

***Supplementary Figure 1. Anti-EBV antibodies do not associate with stroke severity.***

Stroke severity, measured by NIHSS, in relation to EBV antibody titres, (A) total IgG against EBNA1, (B) total IgG against VCA, and (C) IgM against VCA. Each data point represents an individual participant. Analysed by linear regression with best-fit line and 95% confidence band plotted.

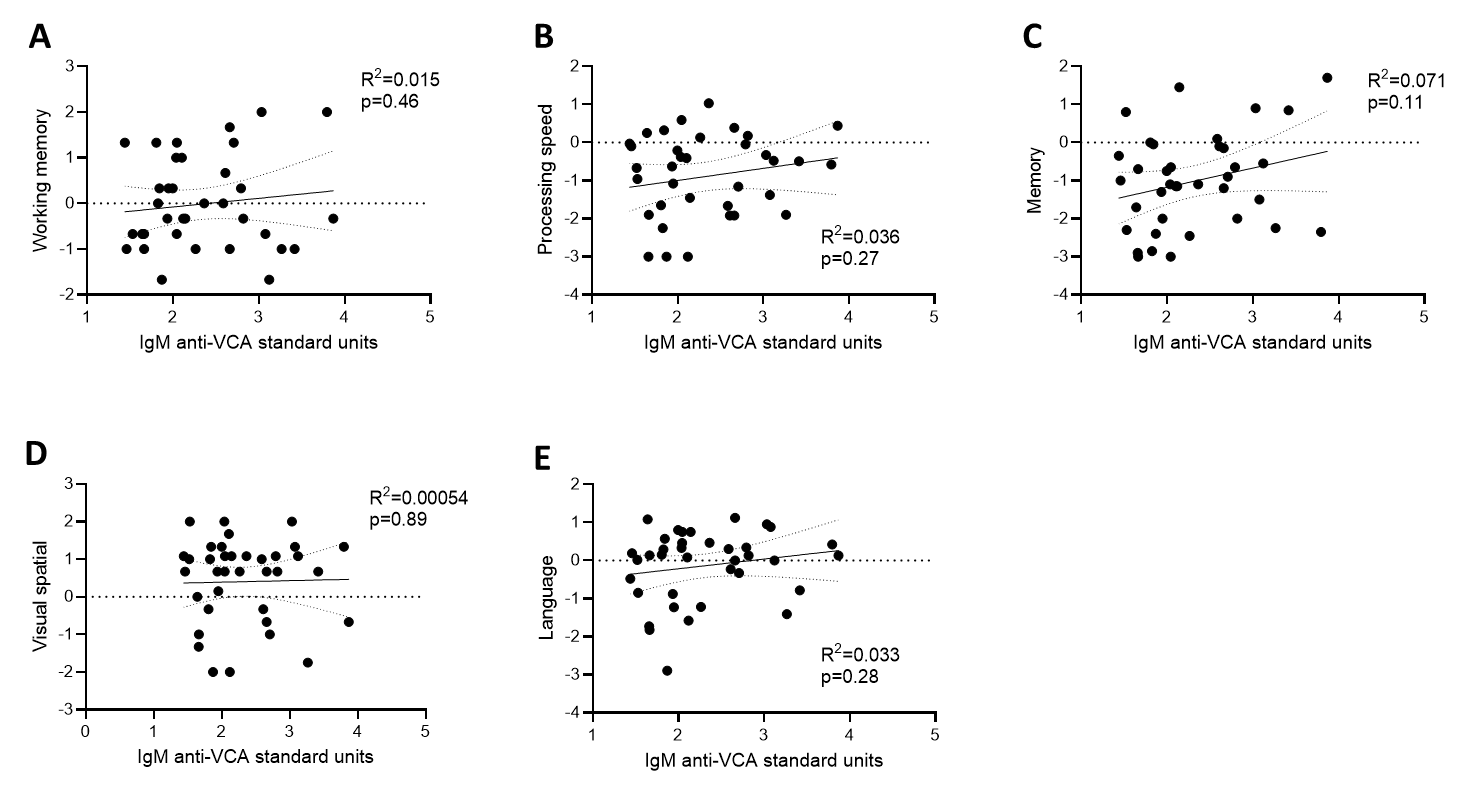

***Supplementary Figure 2. IgM anti-VCA titre does not associate with various cognitive domains***

Titre of IgM against VCA in relation to various cognitive domains, (A) working memory, (B) processing speed, (C) memory, (D) visual spatial, (E) language. Z-score. Cognitive domains are shown as z-scores, or the number of standard deviations above or below the mean. Each data point represents an individual participant. Analysed by linear regression with best-fit line and 95% confidence band plotted.
